# Motivational Interviewing Interventions for Harmful Substance Use Amongst Adults in Low- and Middle-Income Countries (LMICs): A Systematic Review

**DOI:** 10.1101/2023.09.15.23295607

**Authors:** Amy S. Adams, Taryn Williams, Dan J. Stein, Goodman Sibeko, Steven Shoptaw, Stephen Rollnick

**Affiliations:** Department of Psychiatry and Neuroscience Institute, University of Cape Town, Cape Town, Republic of South Africa; Department of Family Medicine, David Geffen School of Medicine, University of California at Los Angeles, Los Angeles, CA, USA; Cardiff University, School of Medicine, UK

**Keywords:** *Substance use disorders Addiction*, *Harmful substance use Motivational interviewing Cognitive behavioural therapy*

## Abstract

**Background:** Harmful substance use is a major challenge globally with far reaching detrimental effects on social, economic and health systems. This burden weighs more heavily on underdeveloped countries as they do not have the infrastructure needed to manage the consequences of substance use. In high-income countries, several evidence-based brief intervention models are used for HSU including Motivational Interviewing (MI). However, fewer studies have been conducted in low-resource settings.

**Aim:** In this review, we systematically reviewed studies utilising motivational interviewing as a component of interventions with adults presenting with harmful substance use in LMICs.

**Methods:** Adhering to Cochrane and PRISMA guidelines, we systematically searched for studies published between 2010 and 2022. The search was conducted in PubMed, EBSCOhost, Web of Science, the Cochrane Library, clinicaltrials.gov and the GSK Clinical Study Register. A narrative analysis was done, and The Joanna Briggs Institute (JBI) Critical Appraisal Tools were used to assess the quality of the studies included.

**Results:** The search yielded 518 publications. Eleven studies from five different countries met inclusion criteria. These consisted mainly of RCTs with some employing qualitative or mixed methods approaches. The focus of the interventions varied, with most targeting alcohol use, while a few addressed opioids, and other drugs. The interventions used different strategies, including MI blended with other interventions (such as CBT, BA, and imaginal desensitization) or using MI-informed approaches. Overall, MI was shown to improve substance use outcomes in seven of the 11 studies with significant reduction in substance use based on outcomes measured.

**Conclusions:** This review highlights that, interventions incorporating MI demonstrated positive effects in improving substance use outcomes in the majority of the studies reviewed. Further research with larger sample sizes and more rigorous study designs is necessary to strengthen the evidence base and address potential sources of bias.

## 1. Introduction

Harmful substance use is a major challenge globally with far reaching detrimental effects on social, economic and health systems [1]. According to the World Health Organization, in 2019 more than 180 000 deaths were directly linked to drug use disorders [2]. The global health risk report states that drug addiction is among the top 20 risk factors for death and disability in the world [3]. Harmful alcohol use alone accounted for an estimated 3.0 million deaths and 131.4 million disability-adjusted life years (DALYs) in 2016, representing 5.3% of all deaths and 5.0% of all DALYs globally [4]. A report by the United Nations Office on Drugs and Crime [5] provides global estimates of substance use prevalence according to substance. According to their report the best estimate for global prevalence of cannabis use is at 4.12%, opioids at 1.21%, cocaine at 0.42%, amphetamines and prescription stimulants at 0.68% and ecstasy at 0.39% [5]. In LMICs the best estimate for prevalence of cannabis use ranges from 2.77-12.00%, opioids from 1.00-3.20%, cocaine from 0.04-2.70%, amphetamines and prescription stimulants from 0.28-1.26% and ecstasy from 0.23-2.84% [5].

Harmful substance use (HSU) is an umbrella term used to refer to licit and illicit substances including alcohol, nicotine, and drugs such as heroin, cocaine, cannabis, amphetamines, methamphetamines and pharmaceutical drugs such as opioids and benzodiazepines [4]. These are substances that produce a psychoactive effect, meaning that when consumed or ingested they affect mental processes including perception, consciousness, cognition or mood and emotions [2]. HSU can therefore be defined as the use of substances that results in any harmful effect on the individual which may include problems in the areas of health, social life, finances, emotional problems, and possibly legal problems [4].

The social burden of HSU is significant and is commonly associated with health concerns, impairment in education and an increasing tendency towards crime and unemployment [6]. This burden weighs more heavily on underdeveloped countries as they do not have the infrastructure needed to manage the consequences of substance use [7]. The World Mental Health Survey showed that 76% to 85% of the patients with severe mental illnesses (e.g., substance use disorders, anxiety or mood) in low- and middle-income countries (LMICs) received no treatment for their conditions over 12 months [8].

In high-income countries, several evidence-based brief intervention models are used for HSU including Motivational Interviewing (MI) alone or with cognitive behavioural therapy (CBT), stress management, problem solving, case management and community contingency therapy [9–11].

However, fewer studies have been conducted in low-resource settings [12]. Multiple factors make delivering effective treatment for HSU challenging in LMICs [13]. These challenges include large geographical spread of health facilities, poorly trained treatment providers to deliver the interventions, stigmatizing attitudes by providers towards HSU and governance that accords low priority for HSU, heavy staff workload [14] poorly resourced health facilities, minimal funding to employ adequate staff among other factors; all contribute towards insufficient services offered to patients presenting with HSU [12].

Motivational interviewing (MI) is a client-centred conversation style counselling approach used to bring about behaviour change [15]. It was initially used to treat individuals with alcohol use disorders but soon expanded to other forms of harmful behaviour and addiction such [16]. At the core of the intervention is a belief in the internal strength and abilities of the individual to overcome their problematic behaviour. It differs from more traditional approaches to substance use interventions which typically make use of direct persuasion, advice-giving or scare tactics to bring about change with limited success [17]. MI instead seeks to elicit an individual’s own internal motivation for change and bring about more sustainable change. It is a type of brief intervention that helps clients identify and overcome ambivalence towards change by utilizing a wide range of therapeutic tools such as open-ended questions, affirmations, reflective listening and summarizing [14].

The effectiveness of MI as an intervention for substance use in HICs is mainly positive but has also produced mixed results. Studies from HICs show that MI results in improved outcomes for non-injection drug users [9] with AUDs [18] improved treatment response [19], for co-occurring alcohol use and depression [20, 21] and has been found to be effective in the treatment of first episode psychosis and SUDs in young adults [22]. However, in a meta-analysis by Wang and colleagues [23] the use of pure MI to treat psychosis and SUDs was found to be varied with relatively modest positive outcomes. Moyers and Houck [24] point out that MI combined with other treatments may at times be challenging as the difference in approaches may be contradictory. They further state that although combined treatment may be effective, they are not always seamless or without difficulty [24].

There is substantially less evidence available for the effectiveness of MI for SUD treatment in LMICs. In a meta-analysis on psychosocial interventions (including three MI-informed interventions) for reducing alcohol consumption in sub-Saharan Africa the outcomes were found to be mixed [25]. The meta-analyses concentrating on AUDIT scores discovered no statistically significant distinctions between the intervention and the comparison group at intervals of 2–3, 6, or 12 months following the intervention. However, reviewing trials related to alcohol abstinence revealed a positive impact of psychosocial interventions compared to the comparison group at the 3–6-month mark post-intervention with sustained effect at longer-term follow-up periods (12–60 months) [25].

Outcomes of intervention research in HICs may not yield similar results in LMICs even after being adapted to the local context in low-income populations [26]. Given the many challenges faced in LMICs it is of utmost importance to identify and implement brief, innovatively delivered, cost-effective solutions that empower providers in primary care settings to effectively treat HSU [12]. This review therefore aims to determine whether HSU interventions incorporating MI is effective with adults in LMICs. This information will be of value to relevant stakeholders working in the field of addiction in LMICs to help make treatment and policy decisions within their given context.

### Aims

In this review, we systematically review studies utilising motivational interviewing as an intervention with adults presenting with harmful substance use in LMICs. In so doing, we aim to determine whether interventions incorporating motivational interviewing as an intervention in this population result in improved clinical outcomes in this setting. We further aim to highlight any limitations within the literature and offer opportunities for future research.

## 2. Methods

Methods were pre-specified and documented in a protocol (PROSPERO– CRD42022329196). The inclusion and exclusion criteria are as follows:

### Inclusion criteria

Studies were included in this review if they met the following criteria:

1. Described an intervention incorporating motivational interviewing, either as a stand-alone or combined with a co-intervention.
2. All randomized controlled trials (RCTs) of interventions incorporating motivational interviewing for harmful substance use and/or substance use disorders (SUD) were included in the review as well as cluster RCTs, RCT feasibility studies, exploratory RCTs, cross-over studies, multiple treatment trials, qualitative studies and studies using mixed methods.
3. Included adult male and/or female participants.
4. Included participants presenting with harmful substance use (as measured by the Alcohol Use Disorders Identification Test (AUDIT), Alcohol Smoking and Substance Involvement Screening Test (ASSIST) or similar) of any substance including alcohol, licit or illicit drugs, tobacco or similar.
5. A diagnosis of an SUD as defined by any operational defined criteria (e.g., DSM-5, ICD-11 or similar).
6. Included participants presenting with or without comorbid mental disorders secondary to SUD (e.g., major depressive disorder, bipolar disorder, generalised anxiety disorder, post-traumatic stress disorder, personality disorders, i.e., borderline personality disorder, antisocial personality disorder, avoidant personality disorder, etc.).
7. Conducted in low- and middle-income countries (LMICs) and settings.

### Exclusion criteria

Studies were excluded based on the following criteria:

1. Where major severe mental illness is present (e.g., Schizophrenia, Bipolar I Disorder, etc), these participants were excluded as these conditions present an additional challenge to the use of motivational interviewing as a treatment approach.
2. Primarily examining healthy populations, children and adolescents.
3. Conducted in high income countries (HICs) and settings.

### Search and data abstraction

The databases searched were PubMed (which includes Medline), EBSCOhost (which includes PsycINFO), Web of Science, and The Cochrane Library. Two trial registries were also searched i.e., clinicaltrials.gov and the GSK Clinical Study Register (2010-2022). The keywords used were all possible combinations of the following words and phrases: “motivational interviewing”, “mi”, “motivational intervention”, “addiction”, “substance use”, “alcohol use”, “drug use”, “tobacco use”, “harmful substance use”, “adults”, “qualitative research”, “quantitative research”, “low- and middle-income countries”, “developing countries”, “developing nations”.

The lead reviewer (AA) systematically evaluated the titles, abstracts and keywords associated with each individual article to determine whether they met inclusion or exclusion criteria. Of the 518 articles found 16 were included. These 16 articles were then reviewed more closely by two authors (AA and TW) and a further nine articles were eliminated. Any confusion or ambiguity about suitability was discussed with the second author (TW) and resolved between the two authors. The reference lists of the remaining seven articles were screened and a further four articles were deemed suitable for inclusion. Therefore, a total of 11 articles were considered suitable for inclusion in this review.

PRISMA guidelines were followed, and the PRISMA search flow diagram was used for this review [27].

Further, two independent reviewers (AA and TW) then extracted the following data: a) description of the trials, b) characteristics of participants, c) characteristics of the intervention, including its duration, d) outcome measures employed (primary and secondary), and e) a summary of continuous and dichotomous data. Additional information, such as the number of total dropouts per group were also extracted. A narrative analysis was then done.

### Assessment of the quality of evidence

Two reviewers (AA and TW) independently assessed the methodological quality of included studies using the Joanna Briggs Institute (JBI) Critical Appraisal Tool for randomized controlled trials (RCTs) [28], qualitative research [29] and analytical cross-sectional studies [30]. The Joanna Briggs checklist for RCTs includes 13 items which covers two domains including *internal validity* (Was true randomization used for assignment of participants to treatment groups? Was allocation to treatment groups concealed? etc.) and *statistical conclusion validity* (Were participants analysed in the groups to which they were randomized? Was appropriate statistical analysis used? etc.) [28]. This appraisal for quantitative evidence is to determine the extent to which a study has addressed the possibility of bias in the area of design, conduct and analysis [28]. The JBI Critical Appraisal Tool for qualitative research includes ten items and considers specific aspects of the study such as congruity between the stated philosophical perspective and the research methodology, representation of participants and their voices and the relationship of conclusions to analysis, or interpretation of the data [29]. The JBI Critical Appraisal Tool for analytical cross-sectional studies includes eight items including whether inclusion criteria were clearly defined, were confounding variables identified and whether outcomes were measured in a valid and reliable way among others [30].

Each study was critically appraised using the relevant appraisal tool and based on analyses the reviewers had the option to include, exclude or seek further information with any disagreement being resolved through discussion between the two reviewers. After independent review all 11 selected studies were included.

### Risk of Bias

Risk of bias was assessed for studies only that met the design. The Cochrane Collaboration ’Risk of Bias’ tool was used, and the following six domains were considered: sequence generation, allocation concealment, blinding of participants and personnel, blinding of outcome assessors, incomplete outcome data, and selective outcome reporting. In addition to this, other sources of bias were also identified and reported on. A judgment on the risk of bias was also made for each domain, based on the following three categories: high risk of bias, low risk of bias and unclear risk of bias [31]. These aspects were covered in the RCT JBI checklist and helped in making final inclusion decisions.

## 3. Results

Figure 1 shows the article selection process of this review according to PRISMA guidelines. The initial searches yielded 518 records, and 495 remained after removing duplicates. After the screening of titles and abstracts, 18 were eligible for full review. Full texts were then reviewed and nine did not meet inclusion criteria. The nine articles identified were excluded based on the following: one article focused on adolescents and not adults; one article was a published protocol and not a full intervention study; one article was an evaluation of a training programme; the study sample in one article was the partner and children of the substance using patient rather than the patient himself; all others were excluded because the study cites were not based in an LMIC. Examining the reference lists from the remaining seven articles identified four additional studies [32–35]. Thus, 11 studies were included in this review [32–42].

**Fig. 1.**
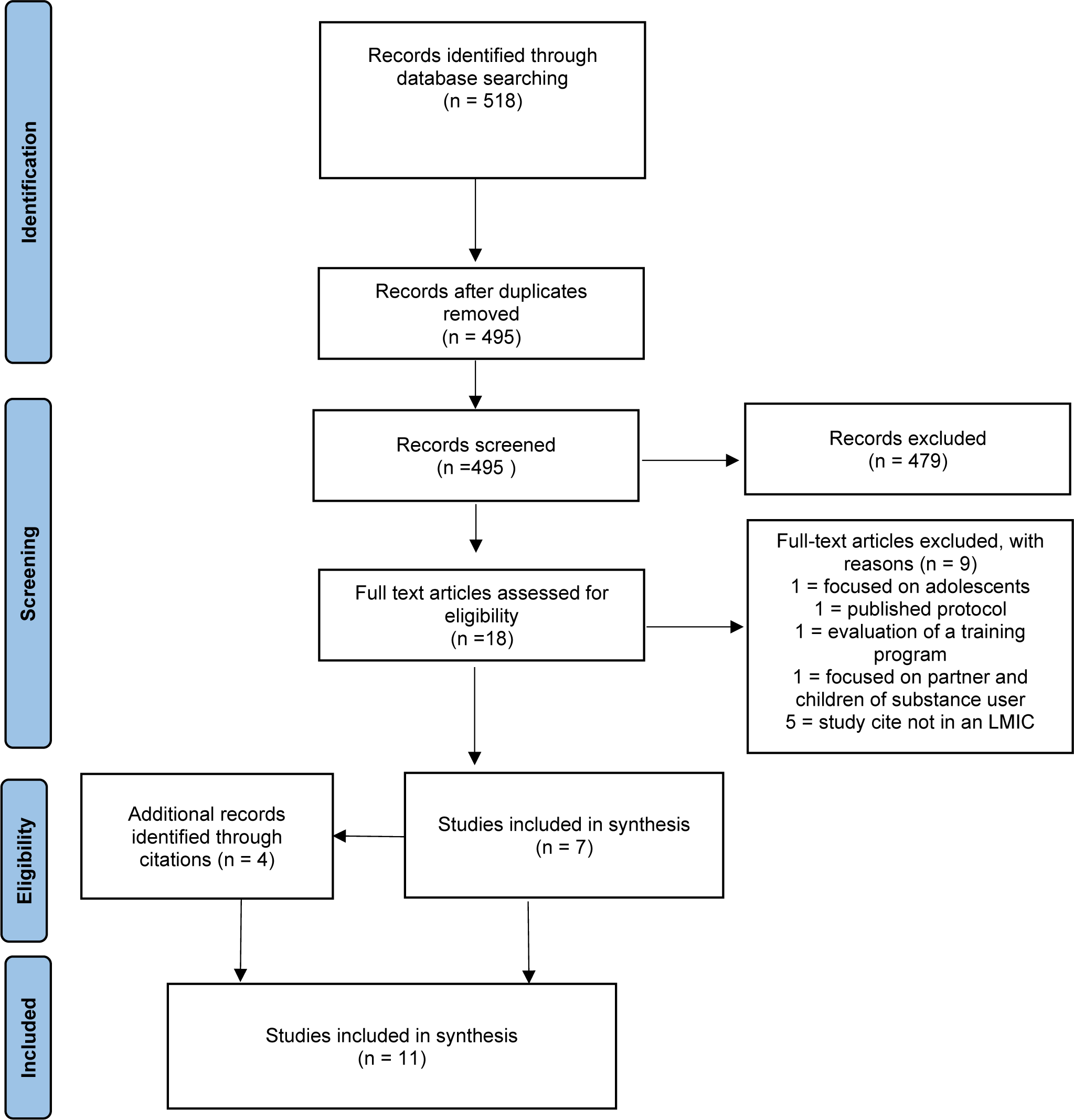
Search flow diagram.

### Trial characteristics Location/setting

As presented in Table 1, four studies were conducted in South Africa [34, 38, 41, 42], two in Zimbabwe [33, 37], two in Kenya [32, 36], one in Uganda [35], and two in India [39, 40]. Study settings included primary healthcare centres [39, 40], emergency centres [42], drop-in centres [32] specialist clinics [33, 35], provincial and district hospitals [37], a residential facility [38], a teaching and referral hospital [36], health care services, and NGOs [41].

**Table 1.** Characteristics of included studies.

| Study | Country | Intervention | Design | Participants | Sample | Mean age | % Male | Key finding/s of outcomes |
| --- | --- | --- | --- | --- | --- | --- | --- | --- |
| Madhombiro et al., 2019 | Zimbabwe (LIC) | MI-CBT: 4 sessions, 30-60 mins per session. | RCT feasibility study | AUDIT scores: Men $\geq 7$ Women $\geq 6$ | N = 40 Dropouts (n = 9) | 39.5 | 57.5% | Significant difference in use over time ($p < 0.001$). No significant difference in the magnitude of change, functioning and QOL. No significant change overtime. |
| Madhombiro et al., 2020 | Zimbabwe (LIC) | MI-CBT: 8-10 sessions, 45-60 mins per session. | Cluster RCT | On ARVs for $\geq 3$ months. AUDIT scores: Men $\geq 7$ Women $\geq 6$ | N = 234 Dropouts (n = 77) | 43.3 | 78.6% | Significant difference in AUDIT score at 6-months post-intervention ($p < 0.001$). |
| Giusto et al., 2022 | Kenya (MIC) | LEAD: BA blended with MI, 5 sessions, 60-90 mins per session. | Qualitative | AUDIT score between 8 and 20 Child between ages 8 and 17 years | N = 9 Dropouts (n = 3) | 39 | 100% | Reduced alcohol use. Improved: mood, interactions at home, feeling of hope, peace in the home, money saving. |
| Marks et al., 2020 | South Africa (MIC) | MI-informed milieu therapy (all staff interactions creating an accepting and positive treatment environment) promoting social cohesion, and elements of restorative justice. | Qualitative | $\geq 12$ months heroin use. ASSIST score: $\geq 27$. Recent use of opioids. | N = 54 Dropouts (n = 24%) | 28 | 96% | High retention rate at 74% and harm reduction. Achieved social cohesion, successfully promoted restorative justice with participants feeling heard, respected, felt a sense of belonging and felt supported. |
| Nadkarni, 2015 | India (MIC) | CAP (MI-informed): 1 to 4 sessions, 30-45 mins per session versus EUC comprising of mhGAP. | Pilot RCT | AUD or AUDIT score of $\geq 19$. | N = 53 Dropouts (n=75.6%) | 41.4 | 100% | Reduced alcohol use in the past 2 weeks but no statistically significant difference between the two treatment groups. Nonsignificant reductions in outcomes between treatment completers and those who dropped out with regard to mean AUDIT (8.6 vs. 12.9, $p = 0.3$), mean alcohol consumed in past 2 weeks (272.5 gms vs. 368.4, $p = 0.6$) and mean SIP score (6.4 vs. 8, $p = 0.7$). |
| Nadkarni, 2019 | India (MIC) | CAP (MI-informed): 1 to 4 sessions, 30-45 mins per session versus EUC comprising of mhGAP. | Exploratory RCT | Attending PHC. AUDIT score of $\geq 20$. | N = 135<br>Dropouts (n = 23) | 43.3 (CAP)<br>39.7 (EUC) | 100% | There was no significant difference between arms for remission at (a) proportion with remission at 3 months (27.1 versus 14.5%, p = 0.18) and 12 months (31.0 versus 18.5%, p = 0.19); (b) proportion of participants reporting no alcohol consumption in the past 14 days at 3 months (35.6 versus 30.7%, p = 0.57) and 12 months (34.5 versus 29.6%, p = 0.57); and (c) consumption among those who reported any drinking in this period at 3 months (58.9 g versus 59.2 g, p = 0.70) and 12 months (45.2 g versus 60.4 g, p = 0.77). For the 'worst case scenario' sensitivity analysis there was no significant difference between the two arms for proportion with remission at 3 months (23.2 versus 13.6%, p = 0.29) and 12 months (26.1 versus 15.2%, p = 0.22). |
| van der Westhuizen et al., 2021 | South Africa (MIC) | One ASSIST-linked MI-informed session followed by two sessions of PST. | Mixed Methods | Attending acute visit at EC. ASSIST scores: $\geq 6$ for alcohol, $\geq 1$ for drugs. | N = 4 847<br>Dropouts = missing information. | 33 | 70% | 59% of patients reported no alcohol use days at the 3-month follow-up, compared to 11% at screening (p = 0.003). Median alcohol scores dropped significantly from the acute EC visit to the 3-month follow-up. Number of drug use days in the previous 2 weeks decreased significantly from screening to follow-up (p < 0.001). At the three-month follow-up, 75% of patients using drugs reported no drug use days in the preceding two weeks. ASSIST scores for patients using cannabis and methamphetamine decreased. |
| Wandera et al., 2017 | Uganda (MIC) | Positive prevention and advice plus a single brief MI-informed alcohol counselling session lasting 20 - 30 mins. | RCT | Continued care at the AIDC for next 6 months. AUDIT-C score of $\geq 3$. | N = 337<br>Dropouts (n = 57) | 39 | 65.6% | Overall reduction in mean AUDIT-C score at 6-month follow-up: 6.4 to 3.4 (p < 0.0001) in the control arm and 6.8 to 3.9 in the intervention arm (p = .001). Mean AUDIT-C score at 3 months was significantly lower in the control arm compared to the intervention arm (3.5 versus 4.3, p = 0.034). No difference in mean scores between groups at 6 months (3.4 in the control group versus 3.9 in the intervention group, p = 0.141). The proportion of participants with mean AUDIT-C scores $\geq 3$ was significantly lower in the control group compared to the interventional arm at 3 months (58.5% in the control group versus 71.5% in the interventional group, p = 0.019) and at 6 months (57.2% in the control group versus 70.2% in the interventional group, p = 0.024). |
| Sorsdahl et al., 2021 | South Africa (MIC) | IDMI: 6 sessions, 20 mins per session. | RCT feasibility trial | MAUD. ≥ grade 9 level of education. | N = 60<br>Dropouts (n = 35) | 31 | 53.3% | Greater reductions in frequency of MA use in treatment group. Statistically significant time by treatment group interaction at both 6 weeks ($p < 0.01$) and 3 months ($p < 0.01$). Consistent with treatment completers at the 3-month follow-up at 6 weeks ($p < 0.01$) and 3 months post-intervention ($p < 0.01$). |
| L'Engle et al., 2014 | Kenya (MIC) | WHO Brief Intervention for Alcohol Use – MI informed, 6 sessions, 20 minutes per session. | RCT | AUDIT scores between 7 and 19. FSWs associated with APHIA. | N = 818<br>Dropouts n = 9% (intervention) n = 7% (control) | 27.5 | 0 | Significant difference in reduction between the intervention and control group on reduced drinking in the last 30 days at 6-month and 12-month follow-up visits, frequency of drinking alcohol, overall binge drinking, binge drinking with paying clients, and binge drinking with nonpaying partners ($p < 0.0001$). |
| Rendall-Mkosi et al., 2013 | South Africa (MIC) | 5 session MI intervention. | RCT | Women at high risk of alcohol-affected pregnancy. | N = 165<br>Dropouts n = 73.39% (intervention) n = 77.11% (control) | 29.8 | 0 | Significant difference in the decline of women at risk for an AEP in the MI group (50.82%) versus the control group (28.12%) at 12 months ($p = 0.009$), 50% reduction in the MI group and a 24.59% reduction in the control group ($P = 0.004$) at the 3-month follow-up. |

*Notes: AIDC = Adult Infectious Diseases Institute Clinic; AEP = Alcohol exposed pregnancy; ASSIST = Alcohol smoking and substance involvement screening test; APHIA = AIDS, Population, Health, and Integrated Assistance II project; AUD = Alcohol use disorder; AUDIT = Alcohol use disorder identification test; AUDIT-C = Alcohol use disorder identification test alcohol consumption questions; BA = Behavioral activation; CAP = Counselling for alcohol problems; EC = Emergency center; EUC = Enhanced usual care; FSW = Female sex worker; Gms = grams; IDMI = Imaginal desensitization plus Motivational Interviewing; LEAD = Learn, Engage, Act, Dedicate; LIC = Low-income country; MAUD = Methamphetamine use disorder; MIC = Middle-income country; MI-CBT = Motivational Interviewing – Cognitive behavioral Therapy; mhGAP = Mental health gap action program; PHC = Primary healthcare center; PST = Problem-solving therapy; QOL = Quality of life; ARVs = Antiretrovirals; RCT = Randomized clinical trial.*

### Participants

Participants were between the ages of 18 and 75 years with sample sizes ranging from nine participants [36] to 4847 [42]. Three studies included men only; of those one focused on fathers with children between the ages of 8 and 17 [36], one with participants presenting with an AUD or AUDIT score ≥ 19 [39], and one with participants attending primary healthcare centres [40]. Two studies included women only; one with female sex workers (FSWs) [32] and the other with women at high risk of alcohol-affected pregnancy [34]. The remaining six studies included both men and women.

One study focused on men and women on ARV treatment [37], one focused on men and women using heroin and/or opioids [38], one study included participants presenting for an acute visit at an emergency centre [42], one study included participants who would continue their care at a specialist clinic for infectious diseases [35], and one study focused on participants with methamphetamine use disorder and participants on HIV treatment [33].

### Design

The included studies were mainly randomized control trials (RCTs) [32–35, 37, 39–41]. Two of the studies used qualitative research methods [36, 38] and one study used a mixed methods approach [42].

### Intervention characteristics

The majority of studies targeted alcohol use [32–37, 39, 40]. The two remaining studies targeted heroin use and opioids [38], alcohol and other drugs [42] and methamphetamine use [41].

### Intervention strategies

All interventions used motivational information, blended motivational interviewing with another type of intervention or were MI-informed. One study used MI only [34]. Two studies combined MI with Cognitive-Behavioural Therapy (CBT) [33, 37]. One study combined MI with Behavioural Activation (BA) [36] and another study combined Imaginal Desensitization with MI (IDMI) [41]. The remaining five studies utilized MI-informed interventions; MI-informed milieux therapy [38]; MI-informed WHO brief intervention for alcohol use [32]; one ASSIST-linked MI-informed session followed by two sessions of PST [42]; positive prevention and advice plus a single brief MI-informed alcohol counselling session [35]; and two studies used CAP (Counselling for Alcohol Problems, an MI-informed intervention) [39, 40]. The number of sessions ranged from one [39, 40] to ten [37], with sessions lasting from 20 [35] to 90 [36] minutes per session.

### Outcome measures

Seven of the 11 studies used the AUDIT as a primary outcome measure to measure the use of alcohol pre- and post-intervention [33–37, 39, 40]. L’Engle et al. [32] initially used the AUDIT but later found that it did not allow for adequate capture of changes in drinking during the study period thus changing their measure to a behavioural interview asking specific questions about drinking behaviour. Of the four remaining studies two used the ASSIST tool to measure the use of heroin and opioids [38], to measure alcohol and other drugs [42] and one study used the timeline follow-back method (TLFB) to measure the number of days participants had used substances within a specified timeframe and the Penn Alcohol Craving Scale (PACS) (modified for MA dependence) to measure MA use and craving [41].

### Impact on substance use

Overall, MI was shown to improve substance use outcomes in seven of the 11 studies with significant reduction in substance use based on outcomes measured. The study by Madhombiro et al. [37] found a significant difference in AUDIT scores at 6-months post-intervention. In their qualitative study, Giusto et al. [36] found reduced alcohol use with improved mood, interactions at home, feelings of hope, peace in the home and money saving in their sample of alcohol using fathers. Marks et al. [38] were able to achieve their goal of a high retention rate of participants at 74% as well as high rates of harm reduction. They also found their intervention had helped them achieve social cohesion, successfully promoting restorative justice with participants feeling heard, respected, feeling a sense of belonging and support [38].

Van der Westhuizen et al. [42] found 59% of patients reported no alcohol use days at the 3-month follow-up, compared to 11% at screening. Moreover, the median alcohol scores had dropped significantly from the acute EC visit to the 3-month follow-up [42]. The number of drug use days in the previous two weeks had decreased significantly from screening to follow-up and at the three-month follow-up, and 75% of patients using drugs reported no drug use days in the preceding two weeks. ASSIST scores for patients using cannabis and methamphetamine had also decreased [42]. Sorsdahl et al. [41] found greater reductions in the frequency of MA use in the treatment group.

Statistically significant time by treatment group interaction at both six weeks and three months. These positive findings were consistent with treatment completers at the 3-month follow-up post-intervention [41].

L’Engle et al. [32] in their study with FSWs observed a statistically significant difference in the reduction of drinking between the intervention and control groups during the 6- and 12-month follow-up visits. Specifically, the intervention group showed a more pronounced reduction in various measures related to alcohol consumption. These measures included a decrease in drinking frequency within the last 30 days, a decrease in overall binge drinking episodes, as well as reductions in binge drinking occurrences involving paying clients and non-paying partners with a p-value less than 0.0001. In a local study [34] a significant difference was found in the decrease of women identified as being at risk for an Alcohol-Exposed Pregnancy (AEP) between the MI group and the control group. At the 12- month mark, the MI group exhibited a decline of 50.82%, whereas the control group’s decline was at 28.12% (p = 0.009). In addition, a 50% reduction was observed in the MI group, compared to a 24.59% reduction in the control group at the 3-month follow-up (p = 0.004). During the 3- and 12- month follow-ups, although not statistically significant, there was a moderate difference in the reduction of risky drinking between the MI group (14.75%) and the control group (10.94%).

Additionally, examining the change in median AUDIT scores from baseline to 12 months, the MI group experienced a more pronounced reduction (a decline of 5 points) compared to the control group (a decline of 1.5 points as determined by Wilcoxon’s rank-sum test, p = 0.007).

The remaining four studies found less favourable outcomes. Wandera et al. [35] report mixed results. They found an overall reduction in the mean AUDIT-C score in the 6 months of follow-up: 6.4 to 3.4 (p < .0001) in the control arm and 6.8 to 3.9 (p = .001) in the intervention arm with the decline in both groups greatest in the first 3 months post-intervention. The mean AUDIT-C score at 3 months was significantly lower in the control arm compared to the intervention arm (3.5 in the control group versus 4.3 in the intervention group, p = 0.034); however, the mean scores were not different between groups at 6 months (3.4 in the control group versus 3.9 in the intervention group, p = 0.141). The only significant finding favourable for MI was shown in a gender-stratified mixed-effects model where the mean difference in AUDIT-C change over time was not different by treatment arm among males (0.38, 95% CI: - 0.41 to 1.17, P = .3493), but among females, the MI arm had greater AUDIT-C reductions compared to the control arm (-1.10, 95% CI: -2.19 to -0.02, p = 0.0457).

Madhombiro et al. [33] found a difference in AUDIT scores over time in both the treatment and control group; the difference between the two groups was however nonsignificant. There was also a nonsignificant difference found in the magnitude of change between the two groups. Nadkarni et al. [39] found no significant difference between arms for remission at three and 12 months, and no alcohol consumption in the past 14 days at three and 12 months. For the ‘worst case scenario’ sensitivity analysis there was no significant difference between the two arms for proportion with remission at three months and 12 months [39]. Nadkarni et al. [40] found that the amount of alcohol consumed in the past two weeks, mean AUDIT score, and alcohol-related problems were all lower in the CAP arm compared to the EUC arm; but the between-group adjusted mean differences were not statistically significant. There were also nonsignificant reductions in outcomes in participants who completed treatment compared with those who dropped out with regard to mean AUDIT scores, mean alcohol consumed in past two weeks, and mean Short Inventory of Problems (SIP) score.

### Risk of bias findings

The risk of bias was assessed for each of the included studies, where applicable (i.e., RCTs: [32–35, 37, 39–41]). Six of the eight RCTs were considered to have low-risk of bias based on the following criteria: having sufficient sequence generation, allocation concealment, blinding of participants and personnel, blinding of outcome assessors, and complete outcome data reported [32, 34, 35, 39].

With regards to the study by Rendall-Mkosi et al. [34] it should be noted that the risk of bias associated with selective outcome reporting was unclear due to the absence of information about a proposal or protocol registration number, which prevents an assessment of whether all intended outcomes were reported as originally planned. The two remaining RCTs were considered high-risk as the blinding of outcome assessors was not reported for both studies [35, 39] for one study [39] the blinding of participants and personnel was not reported and for the other [35] based on what was reported it was unclear whether personnel had been blinded during the intervention phase of the study. High-risk of bias with regards to these aspects of the research means one should be cautious when interpreting results across findings.

## 4. Discussion

The impact of MI as a strategy for addressing substance use in HICs is generally favourable, although it has yielded varied outcomes [9, 18–24]. However, there is far less evidence to support its effectiveness in LMICs. Results of intervention studies conducted in HICs might not demonstrate comparable outcomes when applied in LMICs, even following adaptation for local contexts [26]. Given the limited resources available in LMICs it is essential to identify brief and effective interventions to treat HSU. The purpose of this review was to systematically review studies using MI (whether blended with an additional intervention or as a stand-alone treatment) with an adult population presenting with HSU in LMICs. The overall aim was to determine the efficacy of MI as a psychotherapeutic intervention in the treatment of HSU in resource-constrained settings.

In this review, MI interventions showed significant reductions in substance use and other associated health benefits in seven of the 11 included studies [32, 34, 36–38, 41, 42]. These findings were consistent with results found in HICs [9, 18–22]. There are various potential reasons why the remaining four studies [33, 35, 39, 40] yielded less favourable outcomes.

The study by Wandera et al. [35] reported mixed results with the only favourable outcome for MI found among female participants. The authors proposed that one reason for their results might be that studies reporting positive outcomes in similar populations included participants whose HIV status was unknown or who were HIV-negative, coming from regions with low HIV prevalence. In contrast, their study focused solely on HIV-positive participants. Furthermore, trials involving HIV-positive participants yielded similar findings of no intervention effect [43–45]. It was also found that in studies incorporating more than one session of MI (as opposed to only a single session in this study), lead to reduced number of drinks as well as reductions in heavy drinking [6, 46]. Sample size may have been another contributing factor with relatively small sample sizes in each of the four studies and authors noted that these small sample sizes may not have provided sufficient statistical power to demonstrate significant treatment differences between intervention and control groups and possibly yielding imprecise effect estimates.

Regarding the interventions themselves, Nadkarni and colleagues [40] suggested that Alcohol Dependence (AD) might necessitate a more intensive psychosocial treatment, implying that a brief intervention like the one applied in their study (CAP) might not be adequate to address the complex cognitive and behavioural processes associated with AD. They further proposed that supplementing the CAP intervention with additional strategies might be more effective in improving outcomes for AD, potentially involving discussions on barriers to care, treatment efficacy, pharmacological treatments, and more intensive approaches like telephone monitoring and collaborative case management [40]. This highlights a criticism of individual-level psychosocial interventions for a problem as complex as HSU or SUD. Sileo et al. [47] in their meta-analysis of psychosocial interventions for alcohol use in sub-Saharan Africa argue that research on alcohol-focused interventions in this region would benefit from the inclusion of more structural-level interventions including the reduction of alcohol outlet density, reduced aggressive alcohol marketing, enhanced alcohol regulation and policy enforcement as these are significant contributors to alcohol consumption in African settings.

Wandera et al. [35] found reductions in alcohol consumption as measured by AUDIT-C scores in both the intervention and control arm of their study implying that both interventions contained partially effective components. The comparator in their study may therefore be considered an active intervention as participants received one-on-one counselling which differs from most other studies that use patient information leaflets only in the control arm [48]. Prior research has shown that the simple act of participation and answering alcohol assessment questions in itself may lead to reduced reporting of drinking behaviour [48]. The authors argue that these effects therefore may have distorted the measured intervention effects and could account for the lack of effect noted in their study [35]. Similarly, in the study by Madhombiro et al. [33] the comparator was the World Health Organization Mental Health Action Programme Intervention Guide (WHO mhGAP IG) which may also be considered an active intervention and may have skewed outcomes.

## 5. Limitations

The limitations of this review stem from the variability in alcohol use measurement methods utilised in the included studies, ranging from the AUDIT, AUDIT-C, ASSIST, questionnaires, interviews and other qualitative measures. While the search approach was systematic, it cannot ensure the identification of all interventions, as publication and language biases may have influenced the results. Moreover, certain interventions might not have been included if they did not explicitly mention motivational interviewing, MI, MI-informed or MI-blended as a primary intervention in their methods, abstracts, keywords, or titles. One of the included studies was a pilot RCT, which may have been less robust in design and intervention content [39]. Furthermore, upon evaluating risk of bias, it was found that most of the studies examined in this review demonstrated low risk. Nonetheless, two studies were identified as having a high risk of bias [35, 39] because they lacked information concerning the blinding of outcome assessors and participants or personnel. Therefore, it is advisable to approach the findings of these studies with caution.

## 6. Conclusions

This review highlights that overall, MI interventions demonstrated positive effects in improving substance use outcomes in the majority of the studies reviewed. Significant reductions in substance use, including alcohol, heroin, opioids, and methamphetamine, were observed in several studies. The reviewed studies employed a range of intervention strategies, including MI blended with other interventions (such as CBT, BA, and imaginal desensitization) or using MI-informed approaches.

Despite the heterogeneity, MI interventions consistently demonstrated potential effectiveness in reducing substance use. The studies were conducted in diverse settings, including primary healthcare centres, emergency centres, specialist clinics, hospitals, and NGOs. This suggests that MI interventions can be implemented effectively across various healthcare settings and different geographic locations. The primary outcome measures varied across the studies, with the AUDIT being the most commonly used measure. While many studies reported significant reductions in substance use based on these measures, some studies showed nonsignificant differences between intervention and control groups. Further research with larger sample sizes and more rigorous study designs is necessary to strengthen the evidence base and address potential sources of bias. Nonetheless, MI can be considered a valuable approach for substance use interventions, complementing existing treatment modalities and promoting positive outcomes.

## Data Availability

All data produced in the present study are available upon reasonable request to the corresponding author.

